# Decision Curve Analysis Explained

**DOI:** 10.1101/2025.08.16.25333820

**Authors:** Javier Arredondo Montero

## Abstract

Decision Curve Analysis (DCA) has emerged as a valuable method for evaluating predictive models, yet its application in research—particularly pediatrics—remains limited. This article serves as a didactic tutorial, providing an applied introduction to DCA and clarifying key concepts that often generate confusion. The advantages of DCA over traditional metrics such as the area under the ROC curve (AUC) are highlighted through a simulated cohort of pediatric patients with suspected appendicitis. Three predictors were compared: a composite clinical score (Pediatric Appendicitis Score, PAS), leukocyte count, and serum sodium. While both PAS and leukocyte count achieved acceptable AUCs, the decision curves revealed substantially different net benefit profiles, demonstrating that a higher AUC does not necessarily translate into superior clinical utility. In contrast, serum sodium, with poor discrimination, consistently failed to provide meaningful benefit across thresholds. Common methodological pitfalls—including overfitting, calibration, and the limitations of dichotomous predictors—are also discussed. By contrasting a strong predictor, a moderate predictor, and a weak biomarker, this tutorial underscores the unique contribution of DCA as a practical framework for moving beyond statistical performance toward clinically meaningful decision support.

## Main Text

In pediatric diagnostic and prognostic research, model performance is often assessed using traditional metrics such as sensitivity, specificity, and the area under the receiver operating characteristic curve (AUC). While these measures quantify discrimination, they provide limited insight into the *clinical utility* of a model—that is, whether using it leads to better decision-making in practice.

This gap is particularly relevant in pediatrics, where decision thresholds may vary widely and interventions often carry age- or context-specific risks. A model may exhibit excellent statistical performance but still be unhelpful—or even harmful—if applied in an inappropriate context. Decision Curve Analysis (DCA) addresses this issue.

This manuscript introduces the core principles of DCA, illustrates its construction and interpretation, and demonstrates its application using real data from pediatric diagnostic research.

### Fundamentals and Formula of Decision Curve Analysis

DCA is a method that estimates the *net benefit* of using a diagnostic or prognostic model at different threshold probabilities [1,2]. The net benefit is calculated as the number of true positives identified by a model, penalized by the number of false positives, and weighted by the relative harms of false-positive and false-negative decisions.

Net benefit is calculated as:

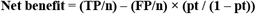

Where *TP* and *FP* are the number of true and false positives, *n* is the total sample size, and *pt* is the threshold probability. This formula reflects the clinical trade-off between the benefits of identifying true positives and the harms of unnecessary treatment. For instance, if the threshold probability is set at 20%, the pt/(1–pt) ratio equals 0.25. This means that each false positive is penalized as one-fourth of a false negative. In practical terms, it would take four unnecessary treatments (false positives) to cancel out the benefit of one correctly treated patient (true positive). This weighting directly reflects the clinician’s tolerance for overtreatment relative to undertreatment.

Suppose a threshold probability of 0.20 is chosen. Then pt / (1 – pt) = 0.25. If a model yields 60 true positives and 40 false positives in a sample of 200 patients, the net benefit would be:

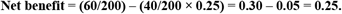

This means the model provides the equivalent benefit of correctly treating 25 additional patients per 100 without unnecessary overtreatment.

The key idea behind DCA is to compare a model not just against chance, but against two clinically relevant extremes: treating all patients as positive or treating none. In the context of a child presenting to the Emergency Department with abdominal pain and suspected acute appendicitis, these strategies would correspond to operating on every child regardless of further assessment (“treat all”) versus discharging all children without additional work-up or surgery (“treat none”). This provides a benchmark to assess whether a model adds value over existing clinical strategies.

By plotting the net benefit of each strategy across a range of threshold probabilities, the decision curve shows whether and when a model provides higher clinical utility than blanket treatment or no treatment at all. A model with a higher net benefit across a relevant threshold range is considered more useful for decision-making.

### Interpreting a Decision Curve

Returning to the previous example of suspected acute appendicitis in children, a decision curve can illustrate the clinical usefulness of a predictive model designed to guide surgical decision-making. The “treat none” strategy assumes that no patient receives the intervention, and by definition its net benefit is always zero across all thresholds. The “treat all” strategy assumes every patient is treated, regardless of their predicted risk. Its net benefit depends on the prevalence of disease and declines as the threshold increases, since a higher threshold penalizes false positives more heavily. Formally, the net benefit of the “treat all” strategy can be expressed as:

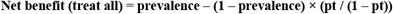

This equation reflects the clinical trade-off of treating every patient—yielding true positives at the rate of disease prevalence, while incurring harm from unnecessary treatment of patients without the disease. As the threshold probability increases, the weight of false positives rises, leading to a progressive reduction in the net benefit of this strategy.

These two reference strategies serve as benchmarks for evaluating the added value of a predictive model.

The x-axis represents the threshold probability—the minimum predicted risk at which an intervention, such as surgery, would be considered. The y-axis shows the net benefit, which combines true positives and false positives into a single metric weighted by the clinical consequences of misclassification.

Typically, three lines are plotted: one for the predictive model, one for the “treat all” strategy, and one for the “treat none” strategy. These serve as benchmarks against which the model’s added value can be judged. The model provides clinical benefit only in the range of thresholds where its curve lies above both reference strategies.

For example, if the model’s net benefit exceeds both “treat all” and “treat none” between 10% and 40% predicted risk, it suggests that its use is preferable to either universal or no intervention within that threshold range. Outside that range, the model may offer no advantage, and simpler decision rules may be equally or more effective.

Decision curves help determine whether, when, and to what extent a model is clinically useful—transforming statistical performance into actionable insight.

### DCA and Prevalence

DCA is influenced by disease prevalence, since the balance between true positives and false positives depends on the frequency of the outcome in the population. In low-prevalence settings, the net benefit of treating all declines rapidly as the threshold increases, while in higher-prevalence settings, treating all may appear favorable over a broader range. Model-based curves are also shaped by prevalence, meaning that the same predictor can show different apparent utility depending on the baseline risk of the cohort. For this reason, decision curves should always be interpreted in light of the population context.

### An Applied Example

To illustrate the application of decision curve analysis (DCA) in pediatric diagnostic research, a synthetic dataset was constructed simulating a cohort of pediatric patients evaluated in the Emergency Department for abdominal pain with suspected acute appendicitis (Supplementary File 1). The dataset comprised 200 simulated pediatric patients, with a prevalence of histologically confirmed appendicitis set at 20% and the remaining 80% representing non-specific abdominal pain. These prevalence values were chosen to approximate conditions in a typical Pediatric Emergency Department and are intended for illustrative purposes. A Pediatric Appendicitis Score (PAS) was calculated for each case. The score ranged from 0 to 10 and was calibrated to approximate the performance reported in external validation studies, where the AUC typically ranges from 0.80 to 0.90 [3].

As comparators, two continuous laboratory variables were included: total leukocyte count, representing a biomarker commonly used in the clinical diagnosis of acute appendicitis, and serum sodium, a marker reported to discriminate between complicated and uncomplicated appendicitis but without value for distinguishing appendicitis from non-surgical abdominal pain [4,5]. The simulated variables were generated to loosely mirror the real-world distribution of clinical findings in pediatric appendicitis, assuming independence between predictors. Each binary variable was assigned a prevalence consistent with existing literature. No correlation structures were imposed between predictors, reflecting a simplified yet educational design.

All statistical analyses were performed using Stata 19.0 (StataCorp LLC, College Station, TX, USA). Logistic regression models were fitted using the Pediatric Appendicitis Score (PAS), leukocyte count, and serum sodium as individual predictors. Discrimination was assessed through the area under the ROC curve (AUC) using 500 stratified bootstrap replications (stratified by outcome status) with percentile-based 95% confidence intervals (seed = 12345). The PAS showed excellent performance (AUC = 0.85; 95% CI: 0.79–0.91), leukocytes demonstrated moderate discrimination (AUC = 0.78; 95% CI: 0.70–0.86), and sodium performed poorly (AUC = 0.64; 95% CI: 0.55– 0.73).

Calibration of all three models was assessed using the *pmcalplot* command (Supplementary File 2). Calibration plots demonstrated good concordance between predicted and observed probabilities for PAS and leukocyte count, whereas serum sodium exhibited systematic miscalibration across the risk spectrum. The Brier score—a metric that evaluates how well predicted risks match actual outcomes, where lower values indicate more accurate and clinically reliable predictions—further confirmed the hierarchy of performance: PAS achieved the lowest mean squared error (0.11), followed by leukocytes (0.13), while sodium performed worst (0.16). These results highlight the superiority of PAS, as it consistently provided the most accurate and clinically meaningful probability estimates.

Decision curve analysis with the *dca* package using a user-written Stata script based on Vickers et al.’s work [6] further highlighted the differences between these predictors (Figure 1). The PAS score demonstrated a consistent and clinically meaningful net benefit across a broad range of threshold probabilities, clearly outperforming both the “treat-all” and “treat-none” strategies, except at very high thresholds (>0.9), where the curve converged with “treat none” as expected given the simulated prevalence. The leukocyte count curve remained above both default strategies up to approximately 0.6, but then fell below “treat none.” This decline reflects the growing penalty for false positives at intermediate thresholds in the context of a 20% prevalence, which outweighs the modest number of true positives contributed by leukocytosis alone. A transient rise was observed between 0.7 and 0.9, driven by a small subgroup of patients with very high leukocyte counts who cross these extreme thresholds and temporarily improve net benefit. However, once the threshold approaches values where virtually no additional cases can be captured, the curve ultimately converges with “treat none.” By contrast, serum sodium did not separate meaningfully from the default strategies at almost any threshold, except for a brief interval between 0.2 and 0.3, underscoring its lack of clinical utility despite an AUC in the modest range (≈0.65). These findings exemplify how ROC-based discrimination can overstate the clinical usefulness of weak predictors when assessed in isolation, whereas DCA provides a more direct evaluation of decision support. Calibration further complements this assessment. In the present example, sodium illustrates how a biomarker can underperform not only in discrimination but also in calibration, underscoring the need to evaluate all three dimensions when judging clinical applicability.

**Figure 1.**
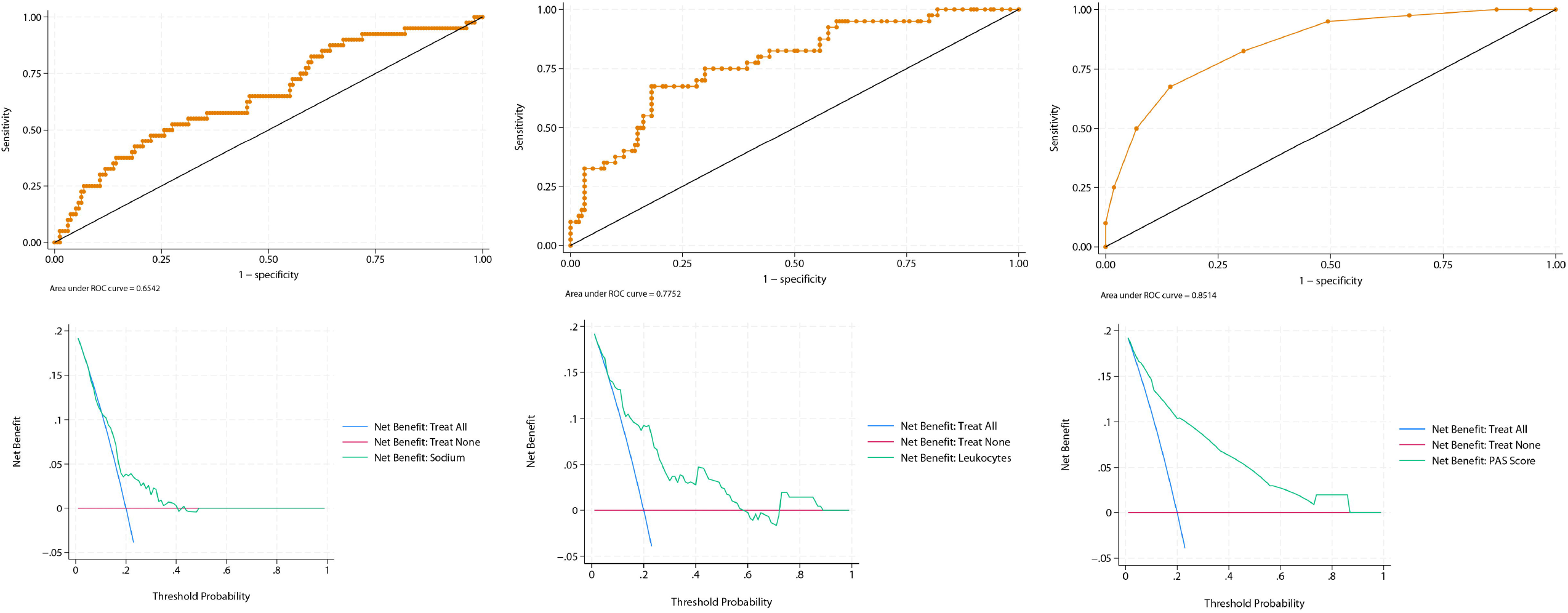
Receiver operating characteristic (ROC, upper panels) and decision curve analysis (DCA, lower panels) for three predictors of acute appendicitis in a simulated pediatric emergency cohort (n=200, prevalence 20%). The left panels display results for serum sodium, which achieved only modest discrimination (AUC 0.64; 95% CI: 0.55– 0.73). Its DCA curve largely overlapped with the “treat-none” and “treat-all” strategies across thresholds, with only a minimal interval of apparent net benefit around 0.2–0.3, underscoring its lack of practical utility despite AUC values commonly reported in the literature. The middle panels correspond to total leukocyte count, which demonstrated moderate discrimination (AUC 0.78; 95% CI: 0.70–0.86). Its decision curve remained above both default strategies up to ~0.6, then declined below “treat none” as false-positive penalties outweighed the modest number of true positives. A transient rise was observed between thresholds of 0.7 and 0.9, reflecting the contribution of a small subgroup with markedly elevated leukocyte counts, before ultimately converging with “treat none” at extreme thresholds. The right panels show the Pediatric Appendicitis Score (PAS), which was calibrated to mirror external validation studies and yielded good overall discrimination (AUC 0.85; 95% CI: 0.79–0.91). In DCA, PAS consistently provided greater net benefit than either “treat-all” or “treat-none” across almost the entire range of thresholds, except beyond 0.9, where convergence with “treat none” occurred as expected given the simulated prevalence. Collectively, these analyses highlight how ROC-based discrimination can overestimate the clinical value of weaker predictors, while DCA offers a more direct appraisal of their decision-making utility.

Two additional DCA analyses were then generated for the PAS score using simulated extreme prevalences within the same dataset, while maintaining a comparable level of diagnostic performance (Figure 2). At a simulated prevalence of 5%, the decision curve showed broad overlap with the “treat none” strategy beyond a threshold probability of 0.5, reflecting the scarcity of true positive cases and the growing penalty of false positives as thresholds increase. Conversely, at a simulated prevalence of 70%, the curve demonstrated consistently high net benefit, exceeding both “treat all” and “treat none” strategies across the entire threshold range. This pattern is expected, as a high baseline risk increases the contribution of true positives and sustains net benefit even at more demanding thresholds.

**Figure 2.**
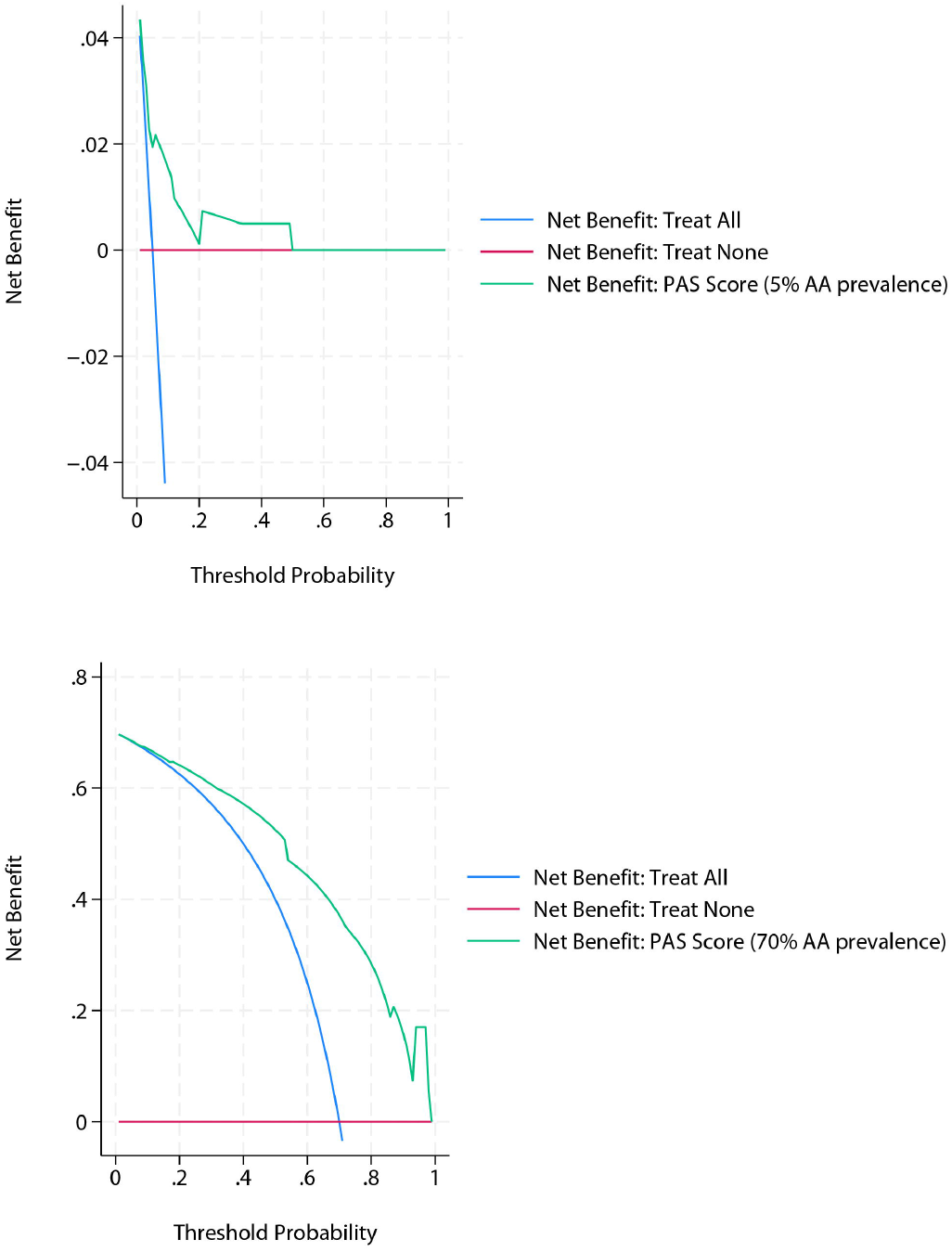
Decision curve analysis of the Pediatric Appendicitis Score (PAS) under simulated extreme prevalence scenarios. At a prevalence of 5%, the curve overlaps substantially with the “treat none” strategy beyond a threshold probability of 0.5, illustrating the limited contribution of true positives and the increasing penalty of false positives in a low-prevalence setting. At a prevalence of 70%, the curve shows consistently high net benefit, remaining above both “treat all” and “treat none” across the full range of thresholds, as expected when baseline risk is high and true positives predominate.

This example illustrates how DCA can reveal significant differences in clinical utility between tools with similar ROC-based discrimination. A score that integrates multiple complementary predictors may offer substantially better decision support than a single biomarker, even when both achieve acceptable AUCs. In pediatric practice, this distinction can directly influence treatment decisions such as observation, imaging, or surgery.

As a final caveat, it should be emphasized that the discrimination, calibration, and decision curve results reported here reflect apparent performance, as they were obtained on the same simulated dataset used for model derivation. In real-world research, external validation or bootstrap correction would be required to avoid optimism and to ensure that the observed net benefit generalizes beyond the training sample

### Strengths and Limitations of DCA

One of the main strengths of Decision Curve Analysis is its ability to incorporate clinical consequences into model evaluation directly [7–9]. Unlike traditional metrics such as AUC or accuracy—which measure statistical performance but ignore the decision context—DCA provides insight into whether a model leads to better outcomes by quantifying the trade-off between true positives and false positives. This makes it especially useful in scenarios with uncertain or variable decision thresholds, where the clinical value of a model depends not only on its discrimination but also on the harm-benefit balance of treatment decisions. DCA also allows models to be compared against real-world strategies such as treating all or none, offering a practical benchmark for clinical adoption.

While DCA is increasingly adopted in diagnostic research, it also has limitations. Its interpretation hinges on the appropriate selection of threshold probabilities, which are often context-specific and subjective. Additionally, the net benefit calculation assumes a uniform misclassification cost across individuals, which may not reflect real clinical heterogeneity. It is also crucial to recognize that DCA does not identify the “optimal” threshold for intervention; rather, it illustrates the net benefit if a given threshold is chosen. In addition, DCA assumes a uniform cost–benefit trade-off across all individuals, an assumption that may not hold in heterogeneous clinical populations. Other complementary methods, such as decision impact curves or net reclassification improvement, may help further characterize model performance. Nonetheless, DCA remains a uniquely intuitive tool for evaluating clinical utility in probabilistic terms. It is also important to emphasize, as highlighted by Kerr et al. [8], that the peak of a net benefit curve should not be interpreted as the ‘optimal’ clinical threshold. DCA is designed to show the relative net benefit across a range of thresholds, not to prescribe a single cut-off for decision-making

### Special Considerations: Overfitting, Binary Predictors, and Calibration

One important methodological caveat in DCA is the risk of overfitting, particularly when the analysis is performed on the same dataset used for model derivation. This can lead to inflated estimates of net benefit that may not generalize to new data. Whenever possible, DCA should be conducted using external validation datasets or bootstrap-corrected predictions to avoid this bias.

Another common issue arises when DCA is applied to dichotomous predictors. Because such variables only generate a few distinct predicted probabilities (often just two), the resulting decision curve reduces to a single straight line segment, connecting the net benefit of the ‘treat none’ strategy to the net benefit observed at one point. This provides information over a much narrower range of thresholds compared with continuous predictors, restricting interpretability and limiting the assessment of clinical Utility.

Finally, DCA assumes that predicted probabilities are well-calibrated. If a model systematically over- or underestimates risk, the net benefit calculation will misrepresent its true clinical utility. Calibration can be evaluated using calibration plots, calibration slope and intercept, or the Brier score. If calibration is poor, techniques like logistic recalibration or shrinkage methods can help adjust predictions and improve decision-analytic performance.

An additional consideration, particularly relevant in pediatrics, is the small sample size often available. Limited data may lead to unstable estimates of net benefit, especially at extreme threshold probabilities. Simulation or bootstrap techniques may be required to assess variability in these contexts.

## Conclusions

Decision Curve Analysis offers a powerful, clinically oriented framework for evaluating prediction models [7–9] (Figure 3). By directly quantifying net benefit across relevant thresholds, it clarifies whether, when, and how a model can improve decision-making. Combined with calibration, it provides a more comprehensive appraisal of predictive performance than ROC analysis alone. In pediatric diagnostics—where both overtreatment and undertreatment carry distinct risks—this dual approach adds value beyond conventional metrics such as sensitivity, specificity, or AUC. For reliable application, however, DCA requires methodological rigor, including adjustment for overfitting and thoughtful handling of predictor types. Its systematic use in diagnostic research can better align statistical evaluation with real-world clinical benefit.

**Figure 3.**
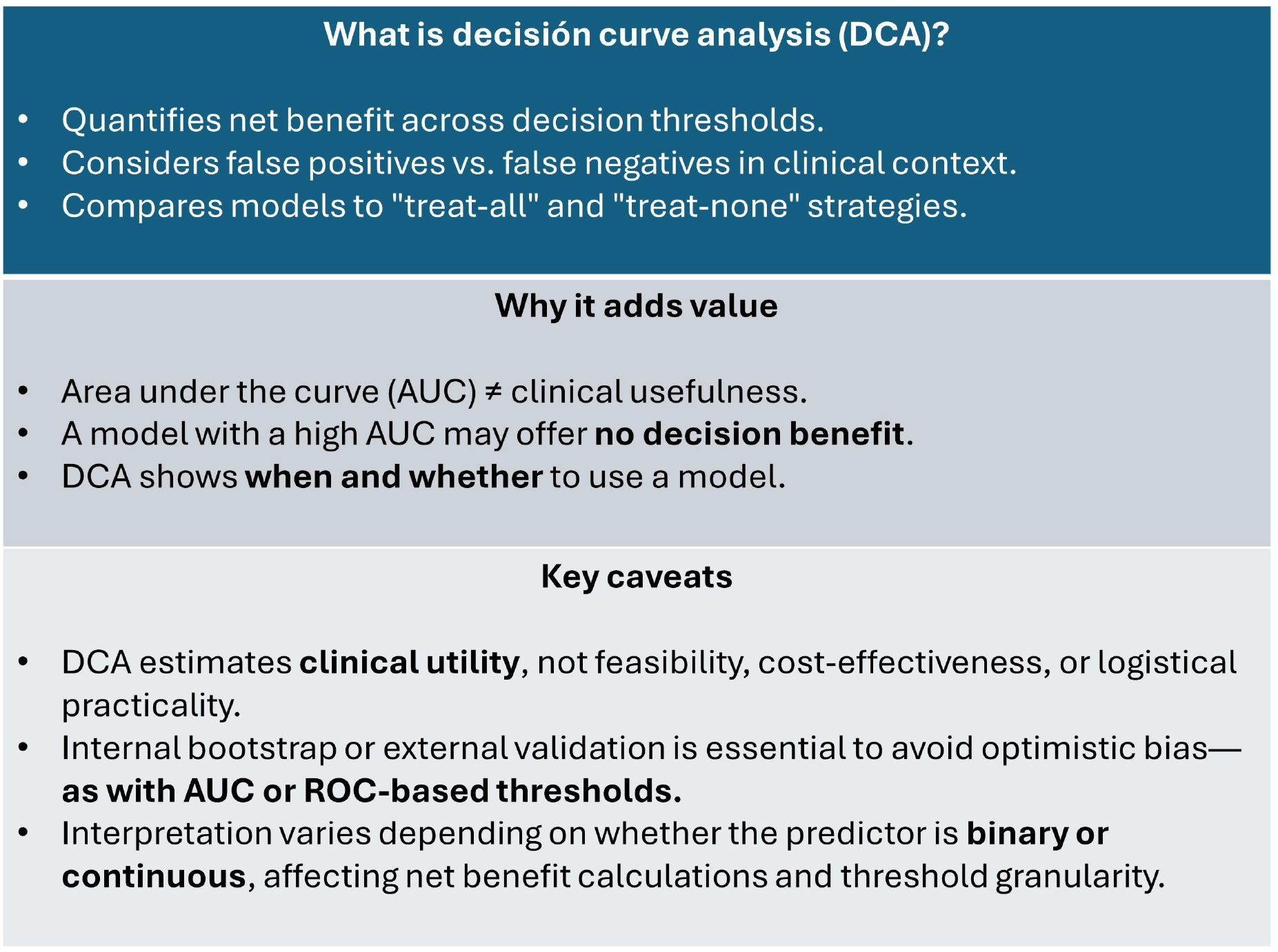
Summary of the core principles of Decision Curve Analysis (DCA). This visual overview outlines what DCA is, how it adds value beyond traditional performance metrics like AUC, and key interpretive caveats. It highlights DCA’s focus on clinical utility—quantifying net benefit across decision thresholds—and its ability to compare models against “treat-all” and “treat-none” strategies. Unlike AUC, DCA shows whether and when a model improves decision-making. However, its interpretation depends on correct threshold specification, external validation, and the nature of the predictor (binary vs. continuous).

## Data Availability

The dataset used in this study is simulated and has been provided as supplementary material (Supplementary File 1).

## Supplementary Files

**Supplementary File 1**. Simulated dataset used for all applied analyses presented in the manuscript. It simulates a cohort of 200 pediatric patients evaluated in the emergency department for abdominal pain with suspected acute appendicitis, including a 20% prevalence of acute appendicitis. The dataset includes a randomized identifier column (id, 1–200), an outcome column for appendicitis (0 = no, 1 = yes), the Pediatric Appendicitis Score (PAS) ranging from 0 to 10 based on its original scale, as well as two continuous quantitative variables: leukocytes (×10^9/L) and serum sodium (mEq/L).

**Supplementary Figure 2. Calibration plots for the three predictors of pediatric appendicitis**. Calibration was assessed using the *pmcalplot* command in Stata. The top panel displays the Pediatric Appendicitis Score (PAS), the middle panel the leukocyte count, and the bottom panel serum sodium. Each plot shows predicted risk (x-axis) against observed outcome probability (y-axis). The 45° line represents perfect calibration. PAS and leukocytes demonstrated good concordance between predicted and observed risks, whereas serum sodium exhibited systematic miscalibration across the risk spectrum.

