## Supplementary figures and images for "Decision Curve Analysis Explained"

### Supplementary File 2

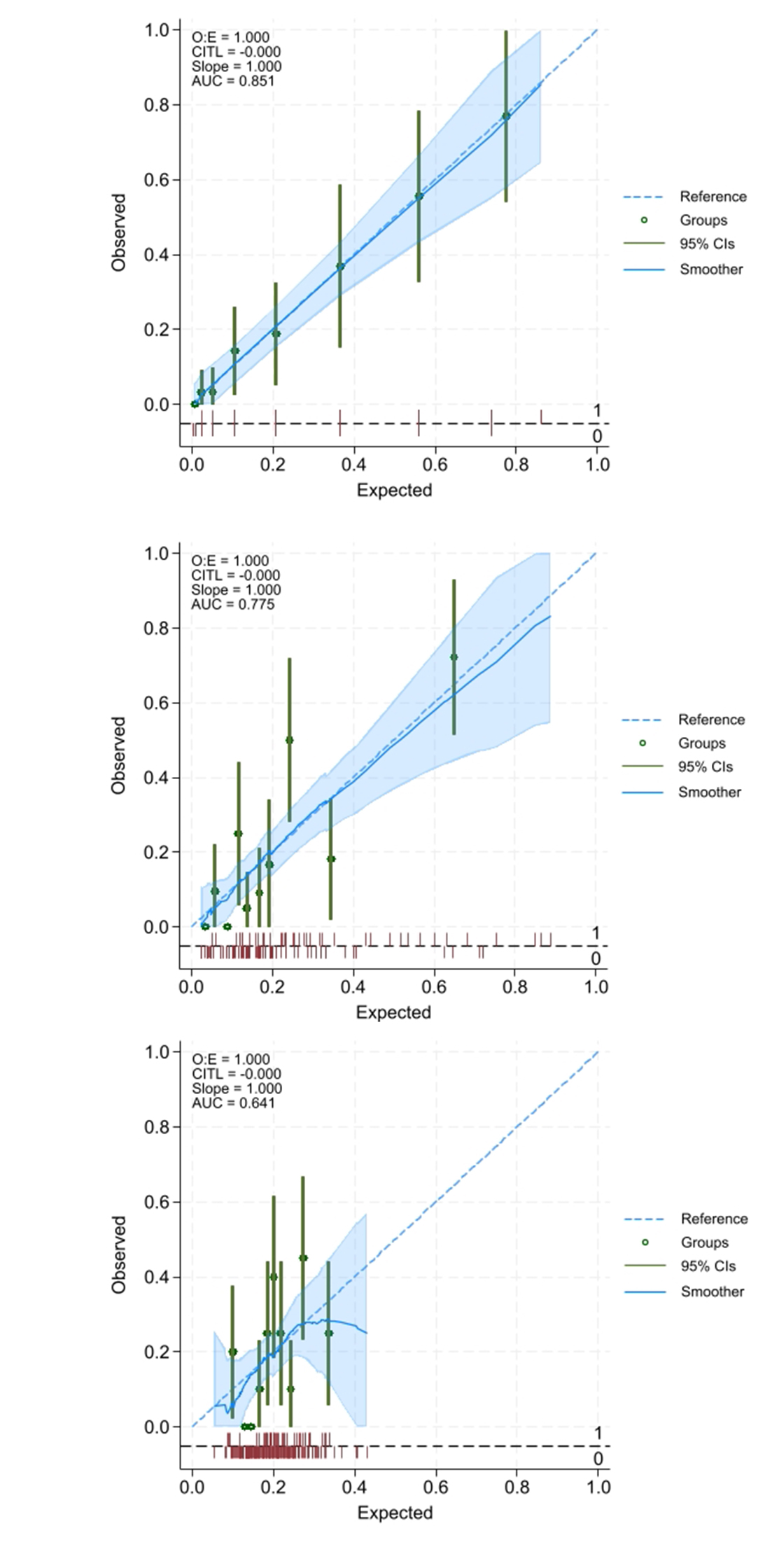
